# Digital Health Technologies for Accessing Contraceptive Services among Young People in Sub-Saharan Africa: A Scoping Review Protocol

**DOI:** 10.1101/2025.01.12.25320439

**Authors:** Agnes Kyamulabi, Eva Oberle, Lotenna Olisaeloka, Innocent Kamya, Ingrid Nyesigire, Abdul-Fatawu Abdulai, Wendy Norman

**Author notes:** Corresponding Author (AK).

## Abstract

**Objective:** This scoping review aims to examine and synthesize existing literature on the use of digital health technologies, with a focus on the extent and types of technologies used to access contraceptive services among young people in Sub-Saharan Africa (SSA).

**Introduction:** Globally, digital health technologies have emerged as pivotal tools in addressing contraceptive needs among young people. In SSA, where traditional healthcare systems often face numerous challenges, these technologies offer innovative solutions to improve access to contraceptive services. Despite growing interest in digital health technologies, comprehensive reviews on contraceptive access among young people in SSA are still lacking. Most existing studies focus broadly on sexual and reproductive health (SRH) or adult populations, leaving a gap in understanding the unique needs and experiences of young people using digital technologies for contraception services. It is unclear how much research has been conducted to examine how these technologies can facilitate contraceptive use, which technologies are used and why, where this evidence is concentrated within SSA, and the prevailing gaps. Therefore, we propose to undertake a scoping review.

**Inclusion criteria:** This scoping review will include studies focusing on young people aged 10-24 years in SSA, addressing access challenges to contraceptive services within this age group. The review will consider client-facing digital health technologies. All methodological approaches and designs will be included. Reviews, protocols, conference papers, policy briefs and studies conducted outside SSA will be excluded.

**Methods:** The review will apply the comprehensive search strategy recommended by JBI. The initial limited search of MEDLINE (Ovid) and CINAHL Complete (EBSCOhost) was conducted with guidance from the University Librarian. This informed the selection of keywords, along with index terms, to develop a full search strategy for MEDLINE (Ovid), CINAHL Complete (EBSCOhost), Scopus, Compendix Engineering Village, and IEEE Xplore. The scoping review shall also use supplementary resources such as google scholar, and African Journal online (AJOL). We will also review the reference lists of articles that meet the inclusion criteria to ascertain articles that were not returned by the search criteria. Data will be presented using tables and charts, accompanied by a narrative summary. This scoping review was registered in Open Science Framework: https://doi.org/10.17605/OSF.IO/5QJ6P

## Introduction

Access to contraceptive services for young people is essential for their physical health, social empowerment, and economic well-being, as it facilitates informed decision-making and prevents unintended pregnancies. The term “young people” refers to adolescents (10-19 years) and youths (15-24 years), as defined by the United Nations (UN).(1,2) Globally, young people constitute about 1.8 billion of the population,(3,4) emphasizing the need for contraceptive initiatives to prepare them for adult life before engaging into child birth. This is because adolescent pregnancy and childbirth come with several complications which are the leading cause of death among girls aged 15-19 years globally.(5) In 2022, an estimated 13% of adolescent girls and young women globally gave birth before the age of 18.(6) Guttmacher Institute reported that each year, around 21 million girls aged 15–19 in developing regions become pregnant, with approximately 12 million giving birth.(7) In 2023, the global adolescent birth rate for women aged 15-19 stood at 41.3 births per 1,000 women, while that for girls aged 10-14 was 1.5 per 1,000.(8) Notably, 10% of all births in 2023 were to adolescent mothers.(8) Despite a general increase in use of modern contraceptives,(8) the proportion of adolescents and young women whose contraceptive needs are met remains lower than that of older women.(7,9) This highlights ongoing challenges in ensuring equitable access to contraceptive services for younger populations.

Adolescent pregnancy significantly contributes to maternal and child mortality(5) and perpetuates the cycle of ill-health and poverty. Despite the numerous risk factors associated with adolescent pregnancies, contraception knowledge and use of modern contraception remain low in many developing nations.(9) The global agenda set by the United Nations’ Sustainable Development Goals (SDGs) for 2030, particularly Goals 1, 3, 4, and 5, emphasizes the importance of enhancing health and promoting gender equality.(8,10) These health-related goals highlight the critical need for equitable access to contraception. Goal 3.7 focuses on achieving universal access to sexual and reproductive healthcare services, including family planning information and education, while integrating reproductive health into national policies by 2030. Regardless of these ambitious plans, a substantial number of adolescents and young women worldwide continue to experience an unmet need for contraception.(7) Unmet need here describes women who do not use modern contraception despite wanting to delay pregnancy or do not want more children.(7,11) Addressing the unmet need for modern contraception among women in low-income regions could significantly reduce the number of unintended pregnancies and unplanned births by 68% annually. (7,12)

Contraceptive use among young people in Sub-Saharan Africa (SSA) is a significant concern due to unique demographic and health challenges. Young people constitute a third of the region’s population,(13) and SSA bears a considerable burden of unintended pregnancies, particularly among adolescents.(12,14,15) High rates of adolescent pregnancies, and unmet needs for contraception are prevalent.(14) In 2023, Sub-Saharan Africa recorded the highest adolescent birth rates in the world, with 97.9 births per 1,000 women aged 15-19 and 4.4 per 1,000 girls aged 10-14.(5,8,9,16) In spite of a global decline in adolescent birth rates—dropping from 64.5 per 1,000 in 2000 to 41.3 in 2023—the region continues to have a disproportionately large number of adolescent pregnancies, with over 6.1 million births to women aged 15-19 occurring in Sub-Saharan Africa.(9,16) Adolescent pregnancies not only disrupt girls’ development but also create lasting barriers to their education, employment prospects, and overall health.(17)

### Digital health for contraception services

In SSA, 68% of adolescents have unmet contraceptive needs. (7) Barriers to contraception include cost, stigma, concerns over potential side effects, lack of knowledge, legal, political, structural barriers, and socio-cultural norms.(7,9,12,18–23) Given the challenges in contraceptive access, digital health is seen as an alternative or complementary mode of providing contraception services to young people. In this review, digital health includes the application of information and communication technology to aid health and health-related areas.(24) The COVID-19 pandemic accelerated the adoption of technology and hastened the transition to digital tools.(25,26) Technology-based and technology-driven interventions have emerged as tools for empowering young people,(25) as they can play a crucial role in overcoming some of the barriers to contraceptive uptake among young people in Sub-Saharan Africa, both directly and indirectly. Given the high burden of unintended pregnancies among young people in Sub-Saharan Africa (SSA) and the transformative potential of digital health technologies, it is crucial to focus on specific interventions that can yield significant benefits, and this is where specifically young people’s access to the technology is important. Digital health technologies, such as mobile health (mHealth) applications, telemedicine, telehealth, e-health, social media and web-based educational resources, offer accessible, confidential, and user-friendly contraception and information services.(27) A qualitative study conducted in higher-income countries revealed that telemedicine for abortion services has the potential to reduce access barriers that were already present before the pandemic.(26) Understanding the most commonly used technologies in contraception service delivery among young people in Africa and their preferences can optimize their deployment(28) and in the long run, they can have a contribution toward reducing unintended pregnancies among young people. For example, mHealth applications have been used to provide contraceptive education, reminders, and connections with healthcare providers.(11,29,30)

Although there is growing interest in digital health technologies, comprehensive reviews examining the types of digital health technologies used, their concentration in Sub-Saharan Africa (SSA), and barriers and facilitators to use are still lacking. Most existing studies focus broadly on sexual and reproductive health (SRH) or adult populations, creating a gap in understanding the unique needs and experiences of young people in SSA who rely on digital tools for contraception services. For instance, Ninsiima et al.(31) examined factors influencing access to youth-friendly SRH services in Africa; Obiezu-Umeh et al. (32) assessed the effectiveness of implementation strategies for youth-friendly SRH in SSA; and Feroz et al. identified a variety of mHealth solutions aimed at enhancing young people’s sexual and reproductive health (SRH) in low- and middle-income countries (LMICs).(33) These studies contribute to a broad understanding of SRH but offer limited insights into how digital technologies specifically contribute to contraceptive access for young people in SSA. Therefore, questions remain on how these technologies facilitate contraceptive use, the types of digital tools used, where the evidence is concentrated, barriers and facilitators and where gaps in knowledge exist. A scoping review is well-suited to address these questions by systematically examining the breadth and nature of evidence on this topic and identifying gaps.

An initial search of Epistemonikos, the Cochrane Database of Systematic Reviews, and MEDLINE confirmed that no reviews on digital contraceptive services for young people in SSA have been registered or conducted. Some systematic reviews have assessed digital interventions aimed at improving contraceptive use, such as Perinpanathan et al.’s review of mobile-based interventions,(11) and Inhae & Jiwon’s meta-analysis of mHealth interventions’ effects on contraceptive use and pregnancy occurrence among young people, focusing only on randomized control trials.(34) However, no review specifically maps the digital health technologies used to facilitate contraceptive services among young people in SSA. Context differences make reviews from high-income countries less applicable, while reviews focusing on LMICs often overlook SSA nations, limiting their relevance. Thus, this scoping review aims to understand the extent and type of digital health technologies used to access contraceptive services among young people in SSA. The review will synthesize evidence on digital technologies facilitating contraceptive services among SSA’s young people, identifying their geographic concentration, exploring associated challenges and opportunities, and highlighting future research needs. Ultimately, findings will inform future research on contraceptive use and unintended pregnancies in SSA’s young population and eventually contribute to improving contraception outcomes.

### Methods

The proposed review will adopt the Joanna Briggs Institute (JBI) methodology for scoping reviews.(35) It will adhere to the PRISMA-ScR (Preferred Reporting Items for Systematic reviews and Meta-Analyses extension for Scoping Reviews) guidelines for reporting,(36) ensuring a comprehensive and transparent approach to mapping the evidence. These guidelines will help to ensure that all key elements of scoping reviews are accurately and clearly presented. This scoping review will identify and map out the various technologies used to access contraceptive services among young people.

### Review question

1. What digital health technologies are currently used to access contraceptive services among young people in Sub-Saharan Africa?
2. Which Sub-Saharan African countries is this evidence concentrated?
3. What are the key opportunities and challenges associated with using digital health technologies to access contraceptive services among young people in Sub-Saharan Africa?
4. What recommendations can be drawn from existing literature to guide future research in leveraging digital health interventions to improve contraception outcomes among young people in Sub-Saharan Africa region?

#### Search strategy

This scoping review will apply the comprehensive three-stage search strategy recommended by JBI(35) to locate both published and unpublished studies. First an initial limited search of MEDLINE (Ovid) and CINAHL Complete (EBSCO) was undertaken to identify articles on the topic. This was done with guidance from University Librarian. Second, the text words contained in the titles and abstracts of relevant articles and the index terms used to describe the articles were used to develop a full search strategy for the different databases to be searched **(see Appendix I**). The search strategy, including all identified keywords and index terms, will be adapted for each included database and/or information source. Third, the reference list of identified reports and articles will be searched for additional sources. This will specifically focus on examining the reference lists of the sources that will be selected from full-text and or included in the review.

The following databases will be searched: MEDLINE (Ovid), CINAHL Complete (EBSCOhost), Scopus, Compendix Engineering Village, and IEEE Xplore. These databases cover a wide range of disciplines, including health sciences and technology. We shall retrieve the search results and export them into Covidence software tool(37) as guided by the librarian to facilitate organization and removal of duplicates. The scoping review shall also use supplementary resources such as google scholar, and African Journal online (AJOL). For Google Scholar, the advanced search feature will be applied, where we will input the same keywords that were used in the Medline (Ovid) search. The search results will then be bulk exported using the Publish or Perish software for further analysis. For African Journal Online (AJOL), we will manually search the platform using the same set of keywords and inclusion criteria to ensure consistency. Relevant studies identified will be individually exported for inclusion in the review. Due to limited resources, only English-language articles and research reports will be considered for inclusion. To avoid bias relating to publication date and to capture the full spectrum of available literature, no date restrictions will be applied.

### Inclusion criteria

#### Population

The scoping review will include papers on digital health interventions providing contraceptive services to young people (aged 10 to 24 years) in Sub-Saharan Africa. This review adopts the UN definition of young people which encompasses adolescents (aged 10 to 19) and youths (aged 15 to 24). The African Union considers a youth or young person to be between the ages of 15-35 years.(13,38) Although this definition represents regional political and socioeconomic realities,(13) this review will utilize the global definition because it captures the important transitional period from childhood to adulthood.(1) This review will include studies focusing on the general population of young people, without specific reference to marginalized or high-risk subgroups (e.g., refugees, sexual minorities, economically disadvantaged youth). Any study that primarily focuses on women of reproductive age (15 – 49) will be excluded.

#### Concept

“Digital health technologies” is the main concept to be explored in this review. Digital health originates from the concept of eHealth, which refers to the application of information and communication technology to aid health and health-related areas.(24) The review will consider studies with a focus on client facing digital health technologies that provide contraceptive services including contraception information, scheduling appointments, consultations, menstruation tracking and links to healthcare services. The review will cover various digital health technologies such as mobile apps, video consultation, telephone calls, text messaging, social media campaigns, and web-based health educational resources. Contraception services are the primary outcome due to the significant access challenges faced by this age group and opportunities presented. Studies addressing contraception outcomes like access, delivery, menstrual health, contraceptive information, family planning counselling, and related topics will also be included. Articles that focus exclusively on service provider-oriented tools, such as electronic health records, digital prescription systems, and billing or insurance platforms, will be excluded. Additionally, studies that primarily address the physical provision of contraceptive services, such as IUD insertion, will not be included. Furthermore, any papers that do not specifically address the use of digital tools for contraceptives or pregnancy prevention but instead focus on other aspects of the broader sexual and reproductive health spectrum, such as STI and HIV/AIDS care, will also be excluded.

#### Context

Studies conducted in Sub-Saharan Africa will be considered and eligible for inclusion if they meet all the other inclusion criteria. World Bank defines SSA as countries that lie south of the Sahara, and these are forty-eight countries.(39) These same countries also make up the World Health Organization African Region.(40) Twenty-four of the twenty-seven countries in the World Bank’s Low-income classification are in SSA. The remaining SSA countries are mostly Lower-Middle income. SSA has the highest youth poverty rate in the world (70%),(41) a factor contributing to the high regional prevalence of teenage or adolescent pregnancy. The review will consider interventions implemented in settings such as schools, universities, health centres and communities within sub-Saharan African countries.

Studies conducted outside SSA or studies that have a combination of countries including those from SSA and those from different continents shall be excluded.

### Types of sources

This scoping review will consider any type of source, without limitations on the methodological approach, including quantitative, qualitative, and mixed-methods study designs. The sources of information will include scholarly journal articles. Research or evaluation reports, dissertations and theses are the only grey literature that will be included in the review. However, reviews, protocols, commentaries, policy briefs, conference papers, and opinion papers will not be included in this review.

### Study selection

Following the search, all identified citations will be uploaded into Covidence software.(37) Duplicates will automatically be removed. A pilot test will be carried out by having two independent reviewers screen at least 5% of the titles and abstracts based on the inclusion criteria. The results will then be discussed, and any necessary adjustments to the inclusion and exclusion criteria will be made in collaboration with the team. After completing the pilot test, two independent reviewers will screen the titles and abstracts of all eligible sources using the inclusion and exclusion criteria. The full text of selected citations will be assessed in detail by two independent reviewers and those that will not meet the inclusion criteria will be excluded and reasons for excluding them will be clearly documented in the final review report. Any disagreement that arises between reviewers at each stage of the selection process will be resolved through a discussion. The results of the search and the study inclusion process will be reported in full in the final scoping review and presented in a Preferred Reporting Items for Systematic Reviews and Meta-Analyses extension for Scoping Review (PRISMA-ScR) flow diagram.(36) In accordance with scoping review guidelines, we will not assess the methodological quality or risk of bias of the studies included in our review.(36,42)

### Data extraction

Data extraction process will be guided by JBI recommendations for data extraction.(43) Data will be extracted by two independent reviewers using a JBI modified data extraction tool(44) **(see Appendix II**). This will also be piloted by 2 independent reviewers using at least 2 papers per source type to ensure consistency; and may be iteratively modified as the review progresses. In situations where additional relevant data items are identified, reviewers will discuss and agree on the items added on the extraction tool and eventually report about the deviation from the protocol and provide the justification.(43) During pilot-testing of the extraction tool the reviewers will reflect on whether there is anything missed on the extraction tool, anything redundant in the extraction tool, whether there is anything in the extraction tool that calls for further clarification, and the time taken to extract the necessary information.(43) All this information will help to guide further the extraction process. The data extracted will include specific details about population characteristics such as country, sample size, age and sex/gender; types of digital health technologies used; contraceptive services accessed using those technologies; study methods and key findings relevant to the review question/s. We shall also extract the title of the papers, authors, objective, evidence concentration, intervention characteristics, key outcomes, gaps and opportunities. Any disagreement that arises between the reviewers at each stage of data extraction process will be resolved through a discussion. If appropriate, authors of the included papers will be contacted to request missing or additional data, where required.

### Data analysis and presentation

For the mapping process, we will create a detailed descriptive map to visually represent key characteristics and the distribution of the studies included in the review. This map will categorize studies based on variables such as population characteristics, geographic location, study design, methodologies used, and key outcomes. To systematically capture this information, we will use a matrix that will allow for the identification of trends, gaps, and the overall scope of the existing literature. This approach aims to point out areas where evidence is concentrated and highlight gaps, thus providing a comprehensive view of the research landscape. We hope to analyse and synthesize the data using basic descriptive statistics such as percentages or proportions; and also apply basic qualitative content analysis, in keeping with the approach to scoping reviews analysis by Pollock *et al*.(43) Two independent reviewers will conduct the basic qualitative content analysis and it will be formed through categorization of the types of technologies; whether the evidence focuses on facilitating access to contraceptive services, challenges and opportunities, and any differences in contraception outcomes; if at all it exists.

A narrative summary will accompany these visual presentations, describing the relationship between the review questions and the findings. Initially, the presentation will detail the authors, publication years, types of sources, settings, study designs, geographic locations, delivery formats, and populations involved in the studies. The second set of data presentation will focus on the digital technologies and applications used to access contraceptive services among young people in Sub-Saharan Africa. It will highlight opportunities the technologies present, discuss challenges faced, and identify gaps in the current research. The presentation will conclude with recommendations for future research and implications for policy and practice.

## Data Availability

This submission is a scoping review protocol and does not involve the collection or analysis of primary data at this stage. As such, there is no pertinent data available for sharing. Once the scoping review is conducted, any relevant data, where applicable and permissible, will be shared in accordance with ethical and legal guidelines.

## Acknowledgements

I acknowledge the contribution of Ursula Ellis for her guidance as a University Librarian and Atul Sharma for his expert advice regarding the relevant existing literature at the initial stages of the study. The proposed scoping review is part of a Master of Science thesis and will contribute to the Master of Science degree for AK.

## Declarations

As an African female student, AK brings both personal and contextual understanding to this scoping review, aiming to synthesize evidence on digital health technologies that support contraceptive services uptake for young people in Sub-Saharan Africa (SSA). Her background contributes a unique perspective on the geographic and social complexities that impact contraceptive access in SSA, enriching the depth of inquiry into existing challenges and opportunities. Through this review, AK strives to foster a broader awareness of the critical need for equitable access to contraception among SSA’s adolescents and contribute to ongoing improvements in contraceptive outcomes across diverse communities in the region.

## Author contributions

AK and WN conceived and designed the study. AK developed the first draft of the study protocol, with critical input from WN, AF, EO, and LO. AK, UE and LO developed the search strategy. AK, IRK and IN did the initial screening to inform stages 2 and 3 of the JBI Search Methodology. All authors reviewed, provided comments, and approved the final manuscript.

## Appendices

### Appendix I: Search strategy

#### Ovid MEDLINE(R) ALL <1946 to July 27, 2024>

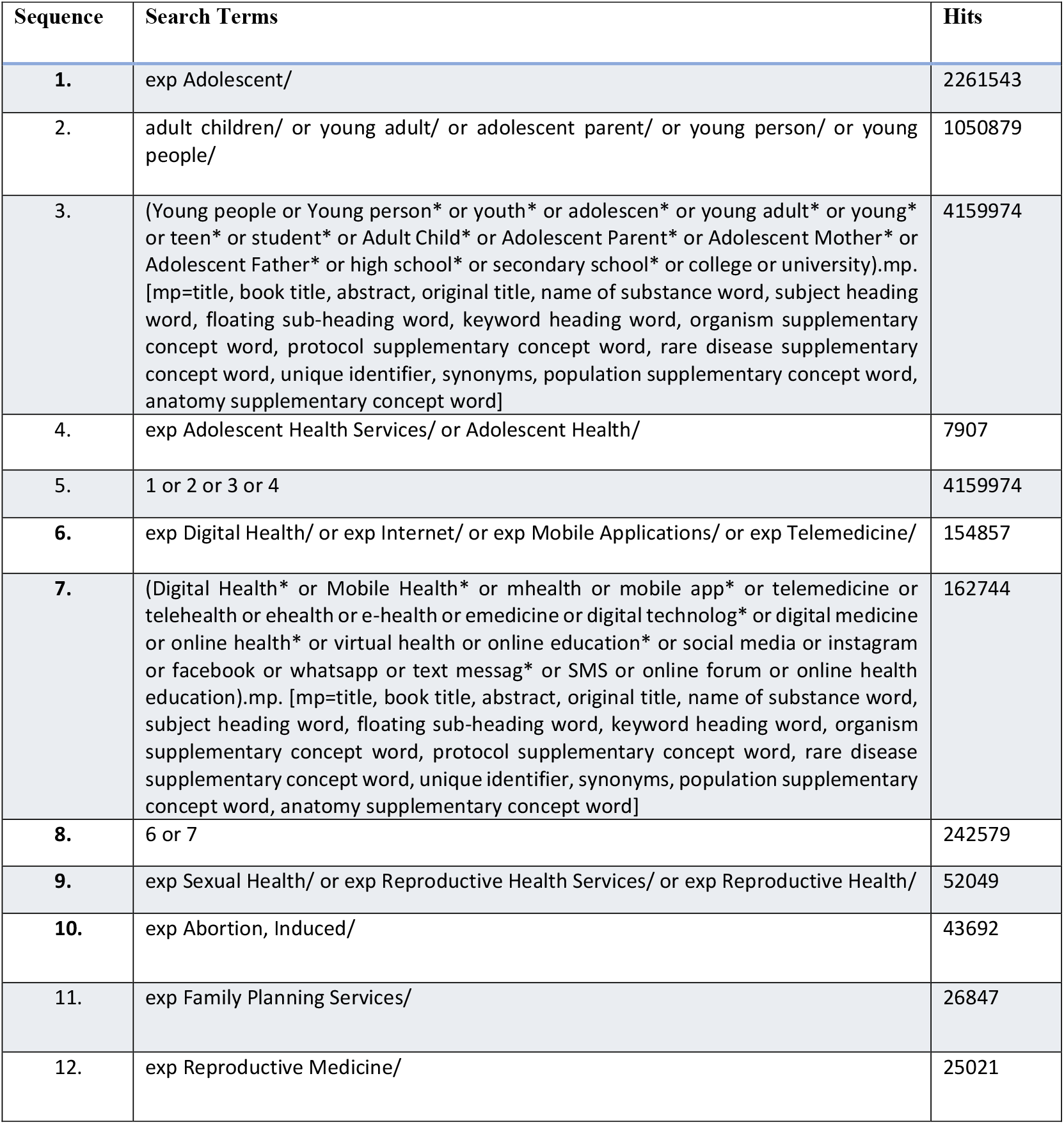
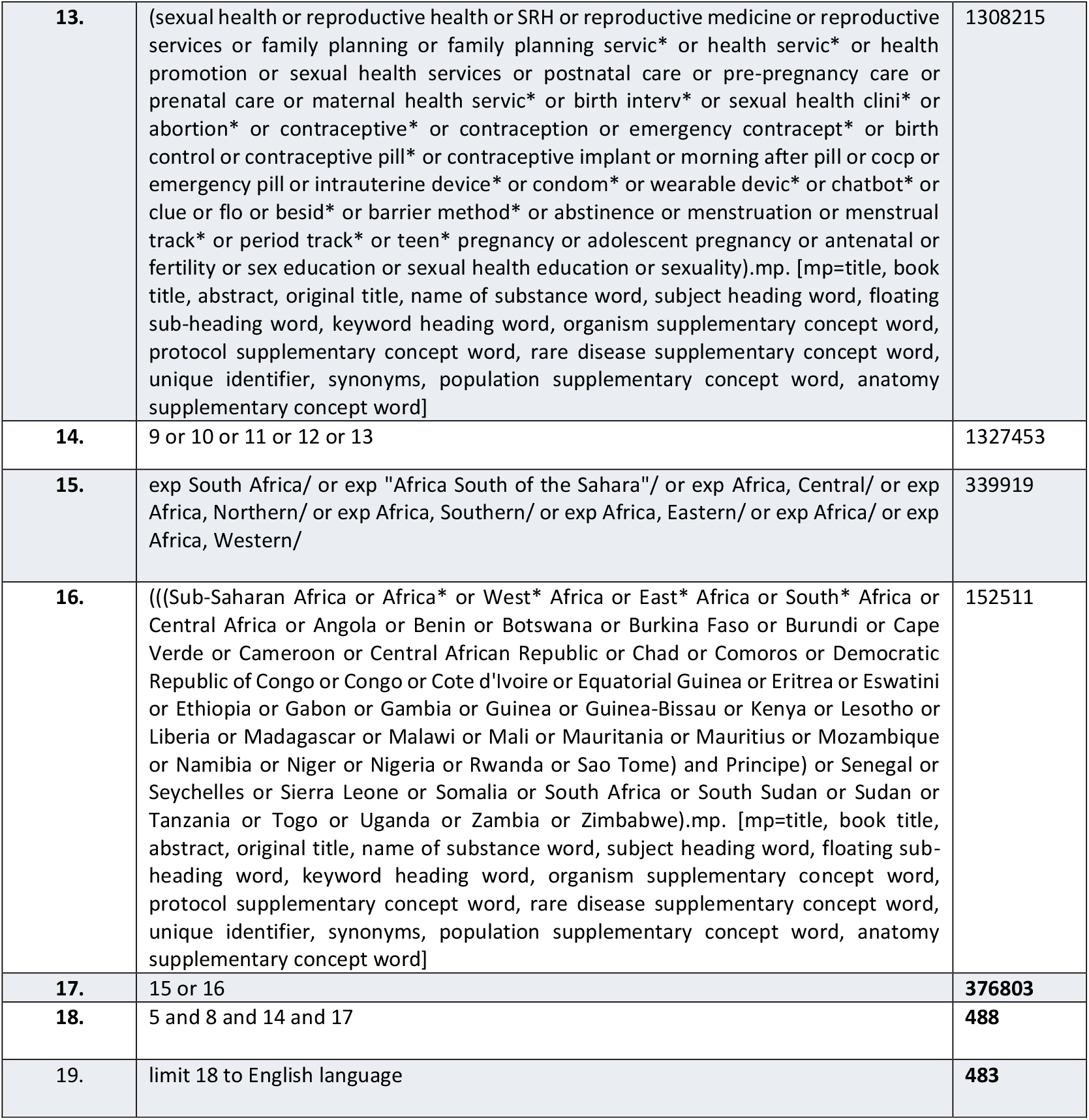

| Sequence | Search Terms | Hits |
| --- | --- | --- |
| 1. | exp Adolescent/ | 2261543 |
| 2. | adult children/ or young adult/ or adolescent parent/ or young person/ or young people/ | 1050879 |
| 3. | (Young people or Young person* or youth* or adolescen* or young adult* or young* or teen* or student* or Adult Child* or Adolescent Parent* or Adolescent Mother* or Adolescent Father* or high school* or secondary school* or college or university).mp. [mp=title, book title, abstract, original title, name of substance word, subject heading word, floating sub-heading word, keyword heading word, organism supplementary concept word, protocol supplementary concept word, rare disease supplementary concept word, unique identifier, synonyms, population supplementary concept word, anatomy supplementary concept word] | 4159974 |
| 4. | exp Adolescent Health Services/ or Adolescent Health/ | 7907 |
| 5. | 1 or 2 or 3 or 4 | 4159974 |
| 6. | exp Digital Health/ or exp Internet/ or exp Mobile Applications/ or exp Telemedicine/ | 154857 |
| 7. | (Digital Health* or Mobile Health* or mhealth or mobile app* or telemedicine or telehealth or ehealth or e-health or emedicine or digital technolog* or digital medicine or online health* or virtual health or online education* or social media or instagram or facebook or whatsapp or text messag* or SMS or online forum or online health education).mp. [mp=title, book title, abstract, original title, name of substance word, subject heading word, floating sub-heading word, keyword heading word, organism supplementary concept word, protocol supplementary concept word, rare disease supplementary concept word, unique identifier, synonyms, population supplementary concept word, anatomy supplementary concept word] | 162744 |
| 8. | 6 or 7 | 242579 |
| 9. | exp Sexual Health/ or exp Reproductive Health Services/ or exp Reproductive Health/ | 52049 |
| 10. | exp Abortion, Induced/ | 43692 |
| 11. | exp Family Planning Services/ | 26847 |
| 12. | exp Reproductive Medicine/ | 25021 |
| **13.** | (sexual health or reproductive health or SRH or reproductive medicine or reproductive services or family planning or family planning servic* or health servic* or health promotion or sexual health services or postnatal care or pre-pregnancy care or prenatal care or maternal health servic* or birth interv* or sexual health clini* or abortion* or contraceptive* or contraception or emergency contracept* or birth control or contraceptive pill* or contraceptive implant or morning after pill or cocp or emergency pill or intrauterine device* or condom* or wearable devic* or chatbot* or clue or flo or besid* or barrier method* or abstinence or menstruation or menstrual track* or period track* or teen* pregnancy or adolescent pregnancy or antenatal or fertility or sex education or sexual health education or sexuality).mp. [mp=title, book title, abstract, original title, name of substance word, subject heading word, floating sub-heading word, keyword heading word, organism supplementary concept word, protocol supplementary concept word, rare disease supplementary concept word, unique identifier, synonyms, population supplementary concept word, anatomy supplementary concept word] | **1308215** |
| **14.** | 9 or 10 or 11 or 12 or 13 | **1327453** |
| **15.** | exp South Africa/ or exp "Africa South of the Sahara"/ or exp Africa, Central/ or exp Africa, Northern/ or exp Africa, Southern/ or exp Africa, Eastern/ or exp Africa/ or exp Africa, Western/ | **339919** |
| **16.** | ((Sub-Saharan Africa or Africa* or West* Africa or East* Africa or South* Africa or Central Africa or Angola or Benin or Botswana or Burkina Faso or Burundi or Cape Verde or Cameroon or Central African Republic or Chad or Comoros or Democratic Republic of Congo or Congo or Cote d'Ivoire or Equatorial Guinea or Eritrea or Eswatini or Ethiopia or Gabon or Gambia or Guinea or Guinea-Bissau or Kenya or Lesotho or Liberia or Madagascar or Malawi or Mali or Mauritania or Mauritius or Mozambique or Namibia or Niger or Nigeria or Rwanda or Sao Tome) and Principe) or Senegal or Seychelles or Sierra Leone or Somalia or South Africa or South Sudan or Sudan or Tanzania or Togo or Uganda or Zambia or Zimbabwe).mp. [mp=title, book title, abstract, original title, name of substance word, subject heading word, floating sub-heading word, keyword heading word, organism supplementary concept word, protocol supplementary concept word, rare disease supplementary concept word, unique identifier, synonyms, population supplementary concept word, anatomy supplementary concept word] | **152511** |
| **17.** | 15 or 16 | **376803** |
| **18.** | 5 and 8 and 14 and 17 | **488** |
| **19.** | limit 18 to English language | **483** |

### Appendix II: Data extraction instrument

#### Scoping review details

Scoping review title:

Review objective:

Review question/s:

#### Inclusion/exclusion criteria

Population

Concept

Context

Type of source

#### Details extracted from source of evidence

1. Author/s
2. Title of the source
3. Year of publication
4. Place of publication
5. Country
6. Objective
7. Study methods/design
8. Population
9. Sample size
10. Age
11. Sex/gender
12. Other demographics
13. Settings
14. Intervention or technology characteristics
15. Contraceptive service provided/received
16. Key findings of the paper
17. Key outcome of the intervention
18. Gaps in the intervention
19. Opportunities presented
20. Gaps in evidence

